# Alpha globin gene deletions in amelioration of clinical severity in beta haemoglobinopathy subjects with the β^0^/β^+^ genotype

**DOI:** 10.1101/2020.05.29.20117135

**Authors:** Dipankar Saha, Prosanto Kumar Chowdhury, Amrita Panja, Debashis Pal, Sharmistha Chakraborty, Kaustav Nayek, Gispati Chakraborty, Prashant Sharma, Reena Das, Surupa Basu, Raghunath Chatterjee, Anupam Basu

## Abstract

Thalassemia is the commonest inherited hemoglobinopathy worldwide. Variation of clinical symptoms entail differences in disease-onset and transfusion requirements. Our objective was to investigate the role of alpha gene deletions in modulating the clinical heterogeneity of thalassemia syndromes. A total of 214 individuals with diagnosed beta-thalassemia major/intermedia were included in the study. Beta globin mutations were determined and categorized as β^+^ and β^0^. Eight common alpha globin gene deletions were detected by multiplex GAP-PCR. Out of the 17 individuals with β^+^/β^+^, 16 did not harbour alpha deletions (αα/αα), and most of them were non-severe. On the other hand, out of 46 individuals with β^0^/β^0^, 30 did not reveal alpha deletions, whereas 16 possessed one or more alpha deletion(s). Accordingly, most of them presented as clinically severe. Out of the 151 β^0^/β^+^ individuals, 119 were negative for alpha deletion, whereas 32 possessed alpha deletions. It was observed that, only in this last category, alpha deletions made a significant contribution (P< 0.0001) in modulation of clinical non severity in this genotype. In conclusion, alpha globin gene deletions play a role to help in ameliorating the phenotype in the β^+^/β^0^ genotype. However, they may have only minor/no role in patients with β^+^/β^+^ or β^0^/β^0^ genotype.

## Introduction

Thalassemia syndromes are inherited monogenic disorders, primarily associated with anemia. Patients typically present with varying degrees of anemia, jaundice, hepatosplenomegaly, bony deformities, growth retardation, endocrine and cardiac dysfunction and failure-to-thrive. Thalassemias display wide variation, not only in the age of disease onset but also with other clinical manifestations. This ailment is caused by the inherited mutations primarily in the beta globin gene that limit the synthesis of hemoglobin [1]. In the *HBB* gene, about 400 different aberrant loci have been identified which are responsible for beta haemoglobinopathies or beta thalassemia [2]. Based on their position and nature they result in major or intermediate forms of beta haemoglobinopathies. β^0^ mutations in *HBB* include defective splicing or early frameshifts, resulting in complete disruption of the mRNA synthesis, hence no beta globin chain is produced. On the other hand, certain substitution mutations may lead to alterative amino acid being placed or mutations in the promoter or Locus Control Region (LCR) may lead to retarded synthesis of the beta chain, hence categorized as β^+^ mutations [3]. However, it has been observed in different populations, that the phenotypic heterogeneity and variations in clinical presentation and manifestations are not always explainable by the mutations solely in the *HBB* gene [4,5]. This leads to diagnostic and therapeutic dilemma.

Several reports are available urging the need to delineate mutations/deletions in the alpha globin genes for clinical management. Most common variations in the alpha globin genes are large deletions of variable lengths. It has been shown that in most cases, despite there being mutations in the alpha globin gene, the subjects remain clinically asymptomatic as there are two copies of the duplicated alpha gene in our genome. The role of alpha mutations as secondary modifiers hence lowering of the disease burden when co-inherited with mutations in the beta globin gene, has been under scrutiny. Several studies from Thailand, Sri Lanka, Bahrain, Central Africa, North America and also India showed the role of alpha globin gene deletions in prediction of disease severity in subjects suffering from beta thalassemia [6–13]. It has also been observed that in several cases of beta thalassemia, presence of alpha mutations may not have distinct clinical effect [14]. It has also been noted by various authors, that, patients with similar types of beta globin gene defects have behaved clinically differently. It is also noteworthy, that till date, about 400 such mutations have been described, but only about 20 of them are responsible for 80% of thalassemia’s [15].

Some patients presenting as beta thalassemia intermedia may have a combination of two known β globin genes with predictive phenotype, but co-inheritance of an α globin gene defect may make it behave differently, as the imbalance in the α/β chain synthesis ratio is lessened. Various authors, tried to explain this mild phenotype with α globin gene mutations, but this model failed to predict the phenotype in its entirety [16,17] However, association of alpha mutations along with beta mutations, remains to be elucidated with proper molecular frame work. Thus, in the present study, it has been investigated in which category of beta globin genotype, deletions of the alpha globin gene may influence the disease severity and helps to build the proper diagnostic protocol for better thalassemia management.

## Materials and methods

### Ethical statement

The present study was approved by the Institutional Ethics Committee of The University of Burdwan, Burdwan, West Bengal. Informed consent and/or assent was taken on case to case basis for participation in this study.

### Subject information

Subjects from both sexes were screened with primary symptoms of hemolytic anemia. Those screened positive for beta haemoglobinopathy by HPLC (homozygous/compound heterozygous states) were selected for this study. Accordingly, 214 subjects had been recruited for the present study. All the subjects were from the West Bengal and surrounding states of eastern India. Clinical and hematological data were collated from the individuals’ medical records. They were classified according to clinical severity or phenotype status as mentioned below.

### Clinical severity or phenotype status

The clinical condition in terms of severe and non-severe had been defined based on the in house method scoring system **[18]**. This was based on age of onset, steady state hemoglobin, transfusion interval, age at first transfusion and spleen status for assessing their clinical condition and to categorize the subjects into “Non-severe” and “Severe” groups.

### Sample collection

Peripheral venous blood samples were collected in K2EDTA observing standard phlebotomy procedure from subjects who were transfused at quarterly intervals or less. Buccal swab samples were collected in 0.9% normal saline (ice-cold) from the subjects who required transfusion at monthly intervals.

### Mutation detection

Mutations in *HBB* gene of the studied subjects were detected by Amplification Refractory Mutation System – Polymerase Chain Reaction (ARMS-PCR), gap PCR and Sanger sequencing. Functional consequence different mutations detected were categorized as β^+^or β^0^, based on the records available in HbVar database [2].

Common deletional forms of alpha globin mutations (HBA), i.e., 3.7kb deletion alpha2 gene (-α^3.7^), 4.2kb deletion alpha2 gene (-α^4.2^), 20.5kb deletion of alpha1 and alpha2 genes (--^20.5^), Southeast Asian deletion of ∼20 kb including alpha1 and alpha2 genes (--^SEA^), South African deletion of 22.8–23.7 kb involving of alpha1 and alpha2 genes (--^SA^), Mediterranean deletion of ∼17.5 kb including of alpha1, alpha2 genes (--^MED^), Thai deletion of 34–38 kb involving the alpha1, alpha2, and zeta genes (--^THAI^) and Filipino deletion of 30–34 kb involving the alpha1, alpha2, and zeta genes (--^FIL^), were determined by the multiplex Gap PCR [19].

### Statistical analysis

To evaluate whether disease severity of different types of beta thalassemia depends on alpha globin gene deletion, Chi-Square (Fisher’s exact) test was performed by using the GraphPad Prism software version 7.0. The significance level was defined at P< 0.05.

## Results

Clinico-pathological and demographic data of the recruited subjects are presented in table 1. Based on the defined scoring criteria, 64 and 150 were classified into clinically non-severe and severe categories, respectively (Table 1).

**Table 1-.** Clinico-Pathological parameters of the studied subjects

| Clinical parameter | Non-severe (n=64) | Severe (n=150) |
| --- | --- | --- |
| Age of presentation, median (range) in years | 6 (0.7 – 40) | 1.92(0.3 - 35) |
| Mean Hb level at detection/ steady state Hb, g/dl (range) | 7.93 (4.5-12.2) | 6.24 (2.1-10.6) |
| HbF % (range) | 27.28 (0.6 – 96.2) | 39.88 (0.4 – 98.6) |
| Prior splenectomy, n(%) | 1 (1.56) | 22 (14.67) |
| Splenomegaly, n(%) | 8 (12.5) | 31 (20.67) |

### Finding of HBB mutations and distributed in

Seventeen different mutations in the *HBB* gene were identified (Supplementary Fig S1 to Fig S2). Among these, 10 mutations were of β^0^ type and 7 mutations were of β^+^ (Table 2).

**Table 2-.**
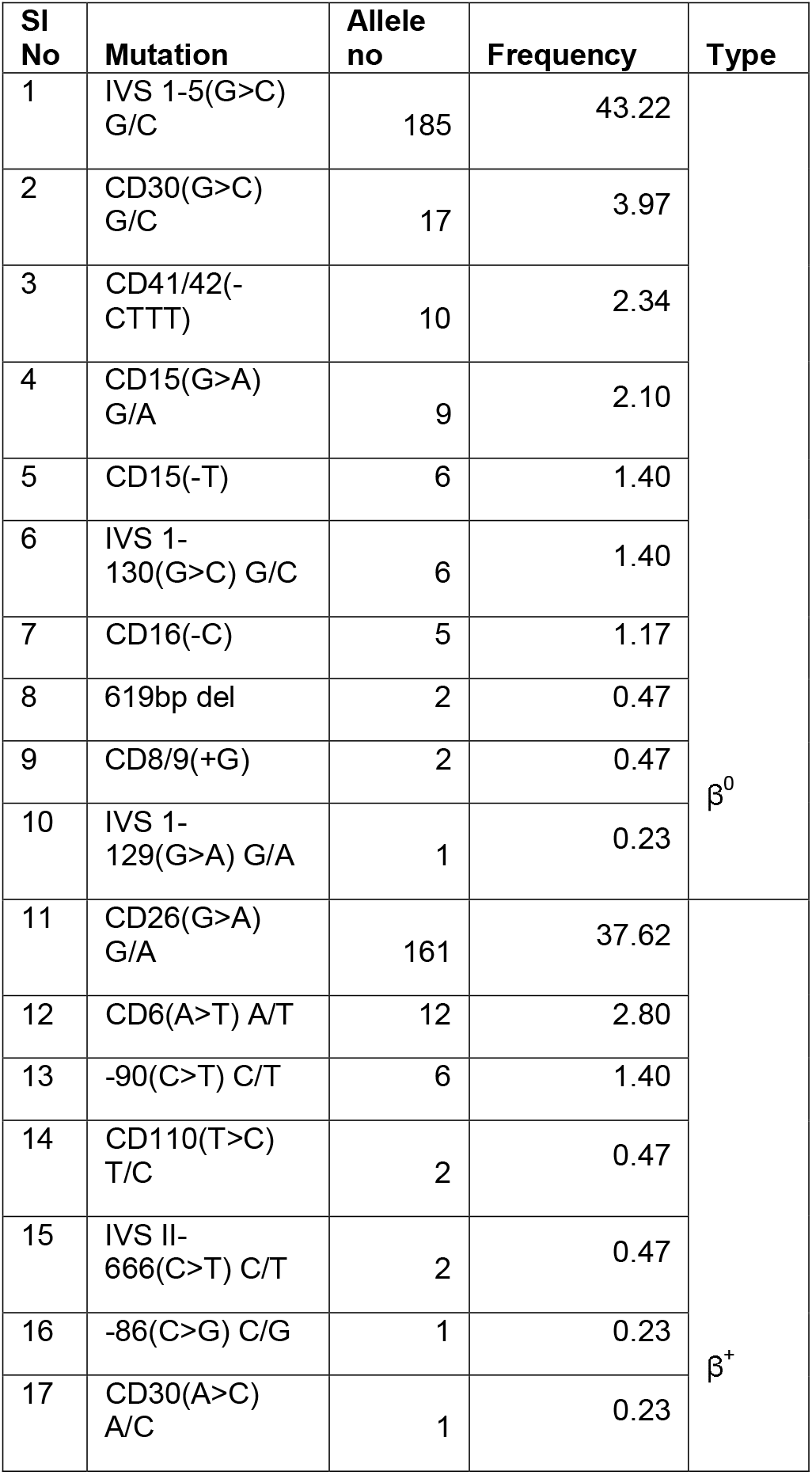
HBB gene mutations and their frequency among studied subjects and their molecular characterization as per HbVar database

Accordingly, the subjects were categorized β^+^/β^+^, β^0^/β^+^ and β^0^/β^0^ based on the *HBB* genotypes. Seventeen subjects were identified as β^+^/β^+^,151 were β^0^/β^+^ and 46 were β^0^/β^0^ (Table 3).

**Table 3-.**
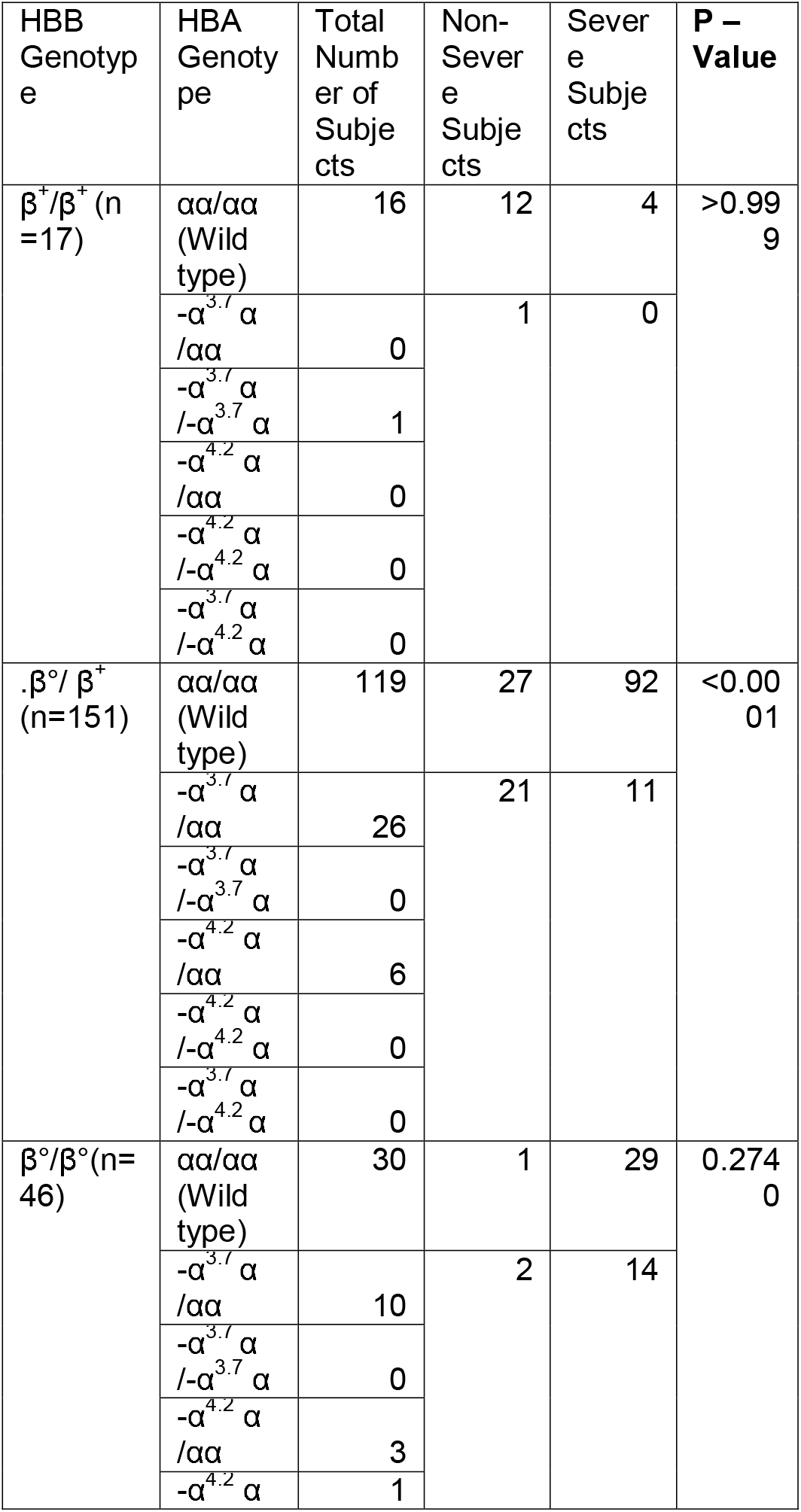
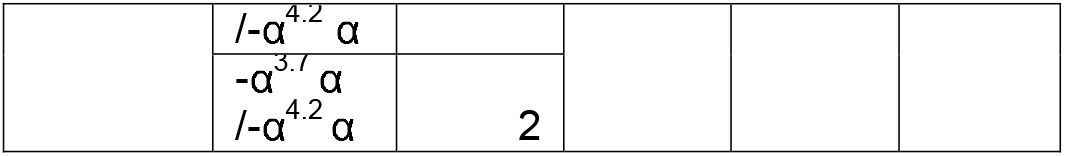
Distribution of subjects with alpha globin deletion mutations among the different beta thalassemia category under Non-Severe and Severe group

### Alpha globin gene deletions

Among the 214 subjects with confirmed homozygous/compound heterozygous *HBB* mutations (either β^+^/β^+^or β^0^/β^+^ or β^0^/β^0^), 49 (22.89%) also harboured one or more deletions in the alpha globin gene (Table 3). Out of the 8 types of deletions screened for, only -α^3.7^ and –α^4.2^ were detected in this cohort in 49 subjects. The remainder (--^20.5^, –-^SEA^, –-^SA^, –-^MED^, –-^THAI^, –-^FIL^) were not detected in any of the patients. 165 subjects had no alpha deletions. Out of the 49 subjects, –α^3.7kb^ deletion was detected in 37 (75.5%) individuals – among these, 36 were heterozygous and 1 was homozygous. Deletion of –α^4.2^ kb was found in 10 (20.4%) subjects, of whom 9 were heterozygous and 1 was homozygous. Two (4.08%) patients were compound heterozygous for both –α^3.7^ and-α^4.2^ deletions [Table 3: Supplementary Fig S3 to S4].

### Effects of alpha globin gene deletions on the clinical severity of *HBB* mutant genotype individuals

Of the 17 individuals with a β^+^/β^+^ genotype, 16 had no alpha deletions (αα/αα), and 12 of these were clinically non-severe (Table 3). On the other hand, out of 46 β^0^/β^0^ genotype individuals, 30 did not have alpha deletions (αα/αα), whereas 16 possessed alpha deletion(s) of any of the types reported in the previous section. Accordingly, a 14 presented as clinically severe. Hence, presence of alpha deletion didn’t make them non severe category. Finally, out of the 151 β^0^/β^+^ individuals, 119 were without alpha deletion (αα/αα), whereas 32 possessed alpha deletions. It was observed that, only in this last category, alpha deletions made a significant contribution (Table 3, P < 0.0001) in modulation of clinical severity in this genotype group, as delineate, in terms of transfusion interval, age of onset, spleen status, etc.[18],

### Discussion

In the present study, 214 subjects with confirmed diagnosis of homozygous/compound heterozygous beta thalassemia were investigated for variations in clinical presentation. Amongst them, 64 were classified into non-severe category and 150 into severe category. Also, to explain the phenotypic heterogeneity, they were categorized by *HBB* genotypes into β^+^/β^+^, β^0^/β^+^ and β^0^/β^0^ types, for revealing the genotypic variation of the primary modifier. To establish the role of alpha deletion in the clinical heterogeneity, they were examined for the presence of common 8 types alpha deletions.

The clinical spectrum of thalassemia patients varies from individual to individual in terms of age of onset, baseline haemoglobin, transfusion interval, spleen size, growth velocity and other clinic-pathological variables. In this study, the parameter of transfusion interval is not taken as sole parameter for severity of disease, rather modified Mahidol scoring system [18] has been followed for consideration of clinical severity of the enrolled subjects, because it considers the cumulative effects of different pathophysiological conditions [20]. It has been observed in the earlier study, person with recurring transfusion may have better overall physiology than person of infrequent transfusion needs. In the present investigation, 6.25% of non-severe category as per Mahidol scoring system needs transfusion of 3 months’ intervals (Table 1)

It is well established that the principal cause of the β-thalassemia syndromes are mutations in the *HBB* gene. It is also well known that the patho-physiological effect of the mutation in the HBB gene depends on the position of the mutation, and criticality of the final gene product, [3]. Depending on the position of the mutation on the HBB gene, the Beta globin protein may be quantitatively and/or qualitatively altered, resulting in no synthesis, sub-normal synthesis or synthesized with altered functional state. It may be qualitatively modified, resulting from substitution of the different amino acid, altering the tertiary or quaternary structure. Hence, affecting its physical and/or bio-chemical characteristics [21]. Thus, in the present study, the identified HBB mutations, have been clustered into β^+^ and β^0^ categories based on their functional and/or beta chain production capacity as per information laid down in the HbVar database. The HBB mutations that results to low synthesis and/or altered function of beta chain have been clustered under β^+^. Those HBB mutations that results in to null or extremely reduced rate of synthesis of beta chain, have been clustered under β^0^. Thus, an individual possessing two copies of β^+^ mutation, i.e., β^+^/β^+^, should have less severe effect as there is chance of Beta chain synthesis with reduced function or at reduced rate. On the other hand, when a person, inherits two copies of β^0^ mutation, i.e., β^0^/β^0^ should face severe mutation effect as there is a little chance of synthesis or function of Beta globin chain of the Hemoglobin. Thus, clinical severity of this two extreme HBB genotype can be explained by HBB mutation itself. Accordingly, in the present study, it has been observed that the majority of the subjects of β^+^/β^+^ is non severe, whereas majority of the subjects β^0^/β^0^ are severe. However, in the subjects of β^+^/β^0^ genotype, as there is chance of synthesis beta chain with or without altered function and clinical condition may not be reflected through the HBB genotype only. In the present study, among 151 subjects, 48 subjects belonged to non-severe category and 103 to severe category. To understand the both type of clinical condition with same type of HBB genotype, in the present study, the role alpha deletions has been hypothesized for further investigation.

The enrolled subjects have been screened for the presence of 8 different types of alpha deletions, which are common in south-east Asian population. Various authors have reported the presence of Alpha gene deletions in Beta thalassemia in different populations [6–13, 20, 22–23].

When we analyzed the extent of deletion in alpha gene, based on heterozygosity (-α/αα) and homo/compound heterozygosity [(--/αα) or (-α/-α)], co-inherited with different types of HBB genotype, it was detected that in the β^+^/β^+^genotype, most of the subjects are non-severe and without alpha deletion. In case of β^0^/β^0^ genotype, most of the subjects are of severe category and without alpha deletion. The number of subjects with alpha deletion is also higher in severe category in this β^0^/β^0^ genotype. Accordingly, it was deduced that there is no significant role of alpha deletion for the modulation clinical severity among the β^+^/β^+^ or β^0^/β^0^ genotype (p< 0.1 and p < 0.27). In contrast, it appeared that Alpha deletion significantly (P < 0.0001) modulated clinical severity amongst the β^+^/β^0^ genotype, where most of the subjects with alpha deletion in this genotype are of non-severe category. Thus, alpha deletions play negligible role in modifying extremes of phenotypes represented as β^0^/β^0^ in the severe group and β^+^/β^+^ in the non-severe group. Accordingly, it could be because that primary defect is so strong that it is enough in its presentation that it does not get significantly altered by the secondary modifier like alpha. Thus, the observations of the present study support the earlier reports that alpha deletion can ameliorate the severity of beta thalassemia, but only in case of β^+^/β^0^ genotype [4,25].

It has been learned that the beta and the non-beta globin chains exist in dynamic equilibrium with each other and strike a critical balance [26]. This balance ensures their mutual stability. If any chain is in relative excess, it precipitates on the inner lining of the erythroid membrane, triggering the hemolytic cascade by stimulating the death receptors, promoting apoptosis. The presence of relative or absolute excess of alpha globin, in beta haemoglobinopathy is the key bio-pathology in causing ineffective erythropoiesis and intra-/extra-vascular hemolysis [3].

Our attempt to explain the clinical phenotype based on alpha deletions yielded non-significant results in both extremes of Non-Severe and Severely affected populations, designated by β^0^/β^0^ and β^+^/β^+^, possibly because the influence required for amelioration is only when a parameter can be synthesized enough to be modified. For example, β^0^/β^0^ individuals may not be synthesizing any beta globin chains at all, so the compensation to adjust the balance would require zero production of the alpha globin chains also. This is physiologically incompatible with life and thus cannot be observed. The observation about the influence of alpha deletions in the equivocal phenotype of β^+^/β^0^ was statistically significant.

Though the observation of the impact of alpha deletions in predicting clinical phenotype was statistically significant in the β^+^/β^0^ phenotype, it could not explain why there was a significant number of individuals with β^+^/β^0^ and 1 individual with β^0^/β^0^ phenotype not harboring alpha deletions still clinically presented as non-severe.

### Conclusion

In summary, subjects having the β^+^/β^0^ mutant allele have one non-functional beta allele and one partially functional allele. The clinical severity among these subjects vary from mild to severe. Accordingly, the alpha globin gene deletion may play a role to compensate the alpha to beta chain ratio and thus help in producing the mild phenotype in the β^+^/β^0^ genotype. Limitation of this study was that the alpha globin triplication and quadruplicating was not checked in the studied subjects. The limitation of the present study can coverup by alpha triplication study and also findings of the modifiers other than alpha

## Data Availability

If requested primary data will be provided

## Conflict of interest

Authors declare there is no conflict of interest in the present study.

## Acknowledgment

The authors are grateful to Department of Science and Technology and Biotechnology, Govt of West Bengal, for supporting this work [Sanc.No-687(Sanc.)/ST/P/S&T/1G-20/2014]. The authors are also thankful to Department of Biotechnology, Govt of India for supporting this work [Sanc. No-BT/PR26461/MED/12/821/2018]. Authors are also acknowledged Burdwan Medical College and Hospital, Burdwan, West Bengal and Institute of Child Health, Kolkata, West Bengal for their support in sample collection and medical data collection.

## Notes

### Competing Interest Statement

The authors have declared no competing interest.

### Author Declarations

Clinical Ethics Committee, The University of Burdwan

